# Differential Causal Effects of Type 1 and Type 2 Diabetes on Osteomyelitis Risk: Insights from Mendelian Randomization Analysis

**DOI:** 10.1101/2024.04.08.24305482

**Authors:** YanHui Li, Chuanyang Zhou, Liming Yang, Lei Tan

## Abstract

**Background:** Osteomyelitis (OM) poses a significant clinical challenge, especially among individuals with diabetes mellitus (DM). While both type 1 diabetes mellitus (T1DM) and type 2 diabetes mellitus (T2DM) have been linked to an elevated risk of OM, the precise causal relationships remain uncertain.

**Methods:** We conducted Mendelian randomization (MR) analyses using summary statistics from genome-wide association studies (GWAS) to explore the causal effects of T1DM, T2DM, their complications, and glycemic traits on OM risk. The study utilized the inverse variance weighted (IVW) method, along with weighted median and MR-Egger for causal estimation, and performed various sensitivity analyses to ensure robustness. Multivariable MR (MVMR) analysis assessed direct effects, while two-step mediation MR analyses investigated the mediating role of DM between rheumatoid arthritis (RA) and OM.

**Results:** The MR analysis unveiled distinct causal effects of T1DM and T2DM on OM risk. Genetically determined T2DM, rather than its complications, significantly increased OM risk (primary dataset: IVW: OR = 1.13, 95% CI 1.056–1.209, *p* = 4E-04; validation dataset: IVW: OR = 1.317, 95% CI 1.14–1.522, *p* =2E-04; Meta-analysis: OR=1.206; 95% CI 1.037–1.402; *p*=0.014), with no observable heterogeneity or horizontal pleiotropy. MVMR analysis confirmed the robustness of the causal association between T2DM and OM, even after adjusting for potential confounders such as body mass index. Conversely, T1DM and its complications showed no significant causal link with OM in either the primary dataset (IVW: *p* = 0.071), the validation dataset (IVW: *p* = 0.276), or the meta-analysis (IVW: *p* = 0.242). Additionally, there was no robust evidence supporting the causal risk of glycemic traits on OM. Mediation MR analysis underscored T2DM as a pivotal contributor to the differential effects of RA on OM.

**Conclusions:** Mendelian randomization analysis provides compelling evidence of a significant causal relationship between genetically determined T2DM and increased OM risk, while T1DM exhibits distinct causal effects. Additionally, our findings highlight the role of T2DM in mediating the association between RA and OM. Further research is warranted to elucidate the underlying mechanisms and guide targeted interventions for OM prevention and management in diabetic populations.

## Introduction

Osteomyelitis (OM), a bone infection, can occur via contiguous spread from surrounding tissues, direct bone trauma, or hematogenous dissemination. It poses a considerable healthcare burden, with a prevalence of 22 cases per 100,000 person-years in the United States, rising over time, especially among the elderly and those with diabetes mellitus (DM)^1^. Challenges in treatment include pathogen identification, bone destruction and repair complexities, and disease recurrence, resulting in prolonged treatment and poorer prognoses^2^.

Diabetes mellitus, affecting nearly 500 million individuals worldwide and projected to increase by 51% by 2045, poses a significant global health challenge^3^. Type 1 diabetes mellitus (T1DM) and type 2 diabetes mellitus (T2DM) are the primary forms, each with distinct pathophysiological mechanisms. While T1DM stems from autoimmune destruction of pancreatic beta cells, T2DM involves insulin resistance and impaired insulin secretion^4^. Although observational studies have linked DM to a higher risk of OM^5,6^, uncertainties persist regarding causal relationships and potential mediators due to confounding factors and biases in existing research. Moreover, limited studies have explored the differing impacts of diabetes subtypes on OM risk.

Clinical studies have established a link between Rheumatoid Arthritis (RA) and heightened susceptibility to OM, attributed to chronic inflammation^7,8^. RA is also associated with diabetes due to a vicious circle perpetuated by glucose derangement and inflammatory mediators.^9,10^. Given the shared risk profile of RA with both diabetes and OM, we employed mediation MR analysis to investigate the mediating role of DM between RA and OM.

Randomized controlled trials (RCTs) are ideal for understanding causality, but their implementation is impractical due to ethical and complex relations between OM and diabetes. MR offers an alternative using genetic variation as a proxy for exposure, mitigating bias^11^. This method mirrors RCTs, validating causal relationships while reducing biases. Moreover, MR elucidates independent causal pathways and potential mediators linking DM and OM.

In this study, we utilized univariable MR (UVMR) analyses to evaluate the impact of T1DM, T2DM, and their complications on OM risk, as well as to explore the association between glycemic traits and OM. Multivariable MR (MVMR) analysis was conducted to assess direct effects by adjusting for potential confounders. Additionally, two-step mediation MR analyses were employed to explore DM’s mediating role between RA, and OM.

## Methods

### 1. Study design and data sources

In this study, we employed UVMR,UVMR and two-step mediation MR analyses to examine the causal effects of T1DM, T2DM, their complications and glycemic traits on OM risk, and to explore DM’s mediating role between RA and OM. Throughout the study, we rigorously adhered to three core assumptions ensured the validity of our results: (1) establishing a reliable association between genetic variants and the risk factor; (2) confirming no association between genetic variants and confounders; and (3) ensuring genetic variants solely influence the outcome through the risk factors. A comprehensive study design flowchart is provided in Figure 1.

**Figure 1.**
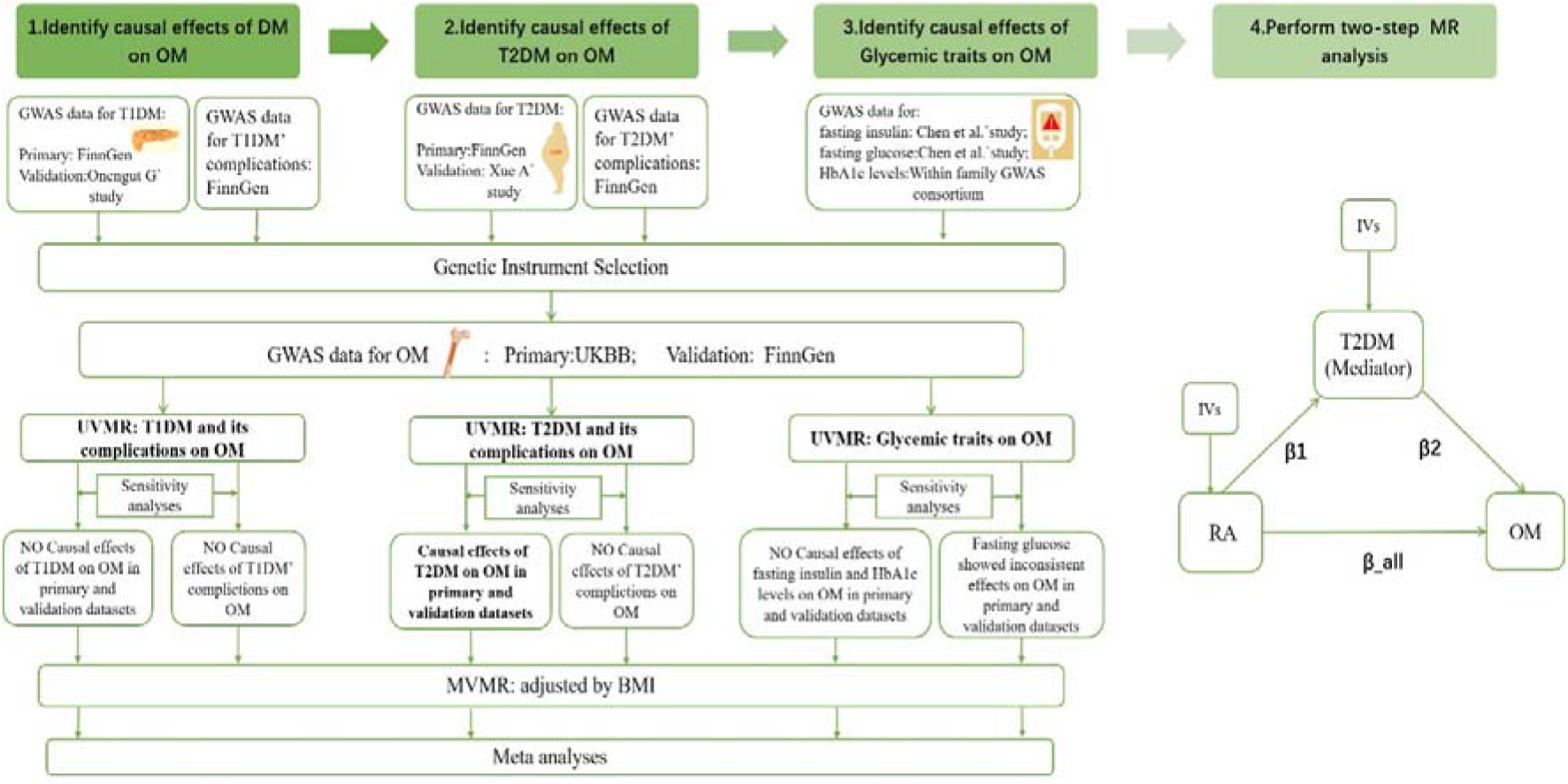
Flowchart of the study design. GWAS, genome-wide association studies; T1DM, type 1 diabetes mellitus; T2DM, type 2 diabetes mellitus; RA, rheumatoid arthritis; OM, osteomyelitis; UVMR, Univariable MR; MVMR, multivariable mendelian randomization; BMI, body mass index; UKBB, United Kingdom Biobank; IVs, instrument variables.

Our study utilized the largest and most recent publicly available summary statistics from multiple genome-wide association studies (GWAS) sources, including the FinnGen database, the UK Biobank (UKBB) database, and other large consortia. To prevent overlap, we carefully selected exposure factors, designating T1DM and T2DM data from FinnGen, and OM data from the UKBB as the primary dataset. DM data from non-FinnGen sources and additional data from FinnGen were designated as the validation dataset. To ensure robustness, we conducted a meta-analysis of results from these datasets. Adherence to the STROBE-MR (Strengthening the Reporting of Mendelian Randomization Studies) guidelines was maintained throughout the study^12^. Detailed information on the GWAS data used in this study is provided in Supplementary Tables 1.

### 2. Genetic association datasets

#### GWAS data for T1DM and T1DM with complications

Type 1 Diabetes (T1DM) genetic instruments were selected from two GWAS Studies. THE Non-UKBB One is based on 5,928 cases and 183,185 controls of European ancestry from the FinnGen database as the primary source. Validation (Non-FinnGen) was conducted meta-analysis using data from Onengut-Gumuscu et al.’s study, which included 6,683 T1DM cases from the UK Genetic Resource Investigating Diabetes cohort and control samples (N=12,173) from four additional cohorts, all reporting European ancestry^13^. T1DM with complications dataset obtained from FinnGen database^14^.Detailed information on single nucleotide polymorphisms (SNPs) used in this study is presented in Supplementary Tables S4 and S6.

#### GWAS data for T2DM and T2DM with complications

The genetic instruments for T2DM were derived from FinnGen database comprising 32,469 cases and 183,185 controls of European ancestry as the primary source (Non-UKBB). For validation (Non-FinnGen), we utilized data from the GWAS meta-analysis study by Xue A et al., which included a sample of 61,714 cases and 593,952 controls with T2DM. This study combined three GWAS datasets of European ancestry, including UKBB, representing the vast majority (99.4%) of individuals of European ancestry^15^. T2DM with complications dataset obtained from FinnGen database^14^. Detailed information regarding the SNPs used can be found in Supplementary Tables S6 and S10.

#### GWAS data for Hyperglycemia traits

SNPs were chosen from a GWAS meta-analysis by Chen et al., involving 151,013 European individuals for fasting insulin, up to 200,622 participants for fasting glucose ^16^. The GWAS summary statistics for HbA1c levels were obtained from Within family GWAS consortium, which included 45,734 participants of European ancestry. All participants had European ancestry, and there was no overlap with the outcome data. Detailed SNP information is available in Supplementary Tables S12, 14 and 16.

#### GWAS data for Body mass index (BMI)

The GWAS summary statistics for MVMR (BMI) were sourced from Within family GWAS consortium, involving 99,998 European participants. There was no overlap between this dataset and other exposures and outcomes considered in our study.

#### GWAS data for RA

The GWAS summary statistics for RA were obtained from Eyre S et al., which included 13,838 cases and 33,742 controls of European ancestry^17^. There was no overlap between this dataset and all exposures and outcomes considered in our study. Supplementary Table 18 provides details of the traits involved in this analysis.

#### GWAS data for Outcomes

The study outcome is OM, defined as inflammation of bone and its structures due to pyogenic bacterial infection. We analyzed the associations between selected instruments and OM using summary GWAS data from the UK Biobank (4,836 cases, 481,648 controls) as the primary source. Validation was performed using FinnGen data (842 cases, 209,575 controls), all of European ancestry.

### 3. Genetic Instrument Selection

For genetic instrument selection, we established a genome-wide significance threshold. SNPs with a P-value less than 5×10^−8^ were considered significant for T1DM, T2DM, glycemic traits, BMI and RA. Since only few SNPs were identified for part of complications of DM when they were as the exposure, a higher cutoff (p□<□1e-6) was chosen. Variants meeting these criteria were then clumped for linkage disequilibrium (LD) using a distance window of 10,000 kB and an r^2^ < 0.01.

To avoid the risk of weak instrumental bias, the F statistic was performed to evaluate the strength of the IV. When F□>□10, the association between the IV and exposures was deemed to be sufficiently robust, thereby safeguarding the results of the MR analysis against potential weak instrumental bias. The PhenoScanner (http://www.phenoscanner.medschl.cam.ac.uk/) was introduced to identify and remove SNPs with potential associations with confounding factors that might violate the independence assumption^18^. After several rounds of rigorous filtering, a set of eligible instrumental variables for the subsequent MR analysis were obtained. Summary of the instrument variables used in this study is presented in Supplementary Tables S2.

### 4. Statistical analysis

We employed the “TwoSampleMR” 22, “MendelianRandomization” 23 and “MR-PRESSO”24 packages for UVMR, MVMR, and Mediation MR analyses, including sensitivity tests. Causal estimates were expressed as odds ratios (ORs) with 95% confidence intervals (CIs). Statistical analyses were conducted using R software version 4.3.2 (The R Foundation for Statistical Computing).

#### UVMR analysis

Causal effects were estimated using the random-effects inverse variance weighted (IVW) method ^19^. To ensure unbiased estimates, MR analyses were also conducted using four alternative methods (MR Egger, Simple mode, Weighted median, and Weighted mode). A causal effect was considered suggested if the IVW p-value was less than 0.05. Moreover, a causal effect was deemed significant if the IVW p-value fell below the Bonferroni-corrected threshold ((p□<□0.05/21□=□0.002) for primary datasets and (p□<□0.05/7□=□0.007) for validation datasets, coupled with consistent directionality in the weighted median and MR-Egger results.

#### MVMR

The DM and BMI shared genetic risk factors^20^. To mitigate the confounding influence of BMI on DM, MVMR analysis adjusting for BMI was performed. For the significant causal associations in the univariable MR analysis, the MVMR analysis was performed using the MVMR-IVW method, aiming to adjust for potential confounding factors BMI^21^.

Mediation MR analysis

Clinical studies indicate that RA is associated with an increased risk of OM^9^. To explore whether DM mediates this association, we conducted two-step mediation MR analysis, assessing three key estimates: (i) the total effect of RA on OM (β_all); (ii) the direct effect of RA on DM (β1); and (iii) the direct effect of DM on OM (β2). Significance was determined by IVW p-values (*p*□<□0.05), with mediation effect proportions estimated using the delta method. The mediation effect is calculated as β1 * β2. The proportion of the mediation effect was estimated as the total causal effect of β_all divided by the mediation effect. We performed two-step mediation MR analyses in both primary and validation datasets to ensure result reliability and compared them for consistency.

#### Sensitivity analyses

Sensitivity analyses were performed to address horizontal pleiotropy and heterogeneity. We utilized weighted median, MR-Egger, and MR-PRESSO methods to verify assumptions and assess robustness, identifying potential horizontal pleiotropy. The weighted median model provides consistent estimates when over half of the weights are from valid IVs^25^. MR-Egger regression detects horizontal pleiotropy and corrects for it, with its intercept term indicating unbalanced directional pleiotropy (*p* < 0.05)^26^. MR-PRESSO identifies and corrects outliers, providing outlier-corrected estimates. The MR-PRESSO distortion test compares estimation differences before and after outlier removal. Cochran’s Q test evaluates heterogeneity among SNPs for exposure and confirms consistency between MR assumptions and analyses (*p* < 0.05)^24^. Credible causal inference requires consistent directionality across the three methods and the absence of horizontal pleiotropic effects.

## Results

### 1. NO Causal effects of T1DM on OM

The causal effect of T1DM on OM was not supported in either the primary dataset (IVW: *p*= 0.071), the validation dataset (IVW: *p =* 0.276), or the meta-analysis (IVW: *p* = 0.242). Sensitivity analyses revealed consistent results with no evidence of horizontal pleiotropy (MR-Egger intercept *p* = 0.521; *p* = 0.967) and no heterogeneity (Cochran’s Q statistic: *p* = 0.089; *p* = 0.244) for the primary and validation datasets, respectively. No outliers were found in the MR-PRESSO test. To further address the potential influence of confounding factors and level pleiotropy, we conducted MVMR. After controlling for BMI, no statistical significance remained between T1DM and OM (Figure 2; Supplementary Table S3). Leave-one-out and scatter plots are provided in Supplementary Figures 1-4.

**Figure. 2.**
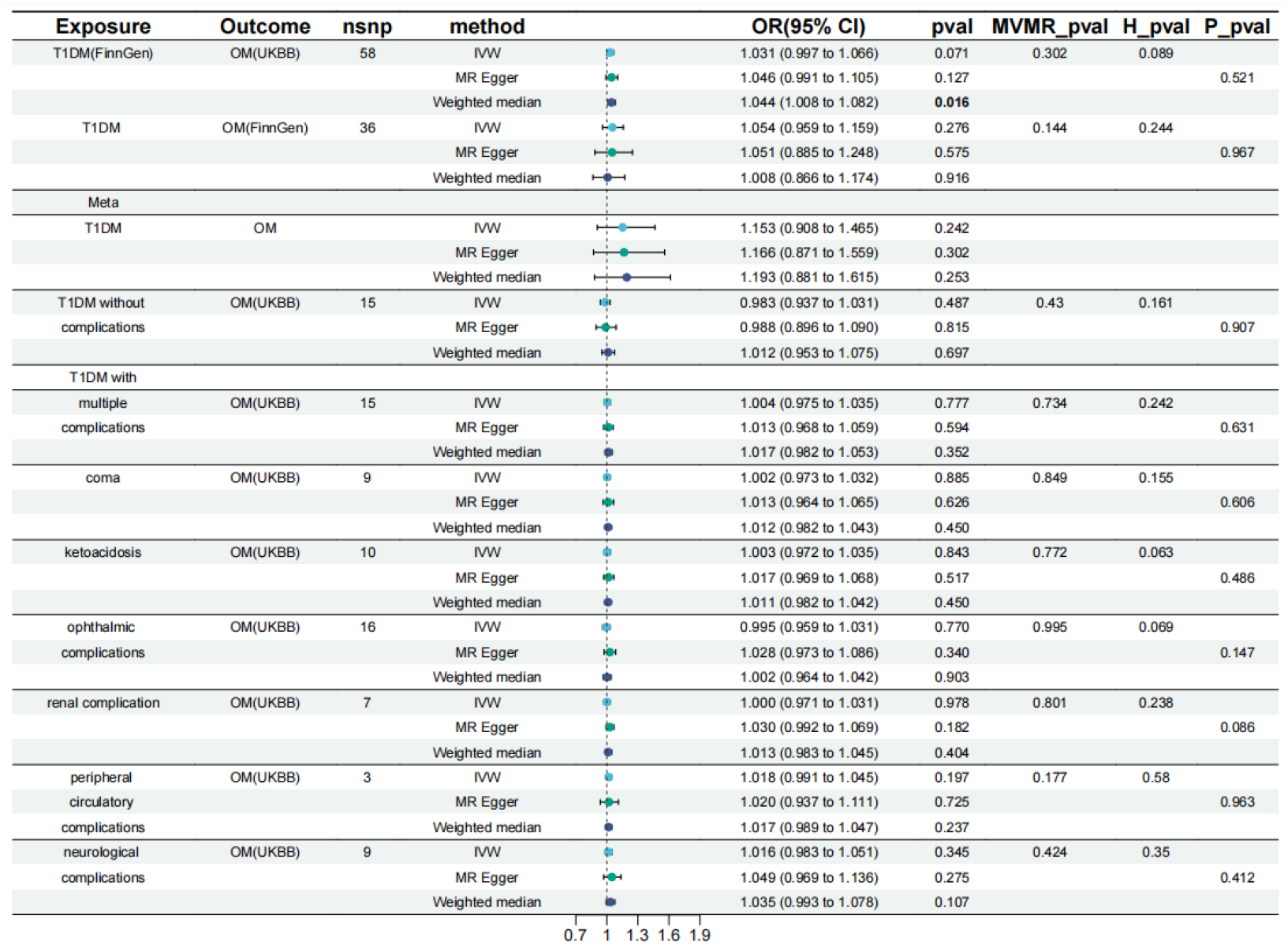
Genetically predicted type 1 diabetes and its complications: associations with osteomyelitis. T1DM, type 1 diabetes mellitus; OM, osteomyelitis; UVMR, Univariable MR; MVMR, multivariable mendelian randomization; OR, odds ratio; CI, confidence interval; UKBB, United Kingdom Biobank; IVW, Inverse Variance Weighted; H, Heterogeneity; P, Pleiotropy. p-values (IVW)□<□0.05 was considered suggested different. The Bonferroni-corrected results of the p-values (IVW) in each group remained consistent with the uncorrected results.

To explore the relationship between different T1DM subgroups and OM, we analyzed data from the comprehensive FinnGen database, known for its coverage of T1DM complications. Our analysis revealed no causal correlations between either T1DM without complications or T1DM with complications and OM (IVW: *p*L=L0.478; *p*L=L0.777, respectively). Further subgroup analyses within T1DM with complications showed no causal links with OM. Specifically, IVW -P values for OM were 0.885 for T1DM with coma, 0.843 for T1DM with ketoacidosis, 0.770 for T1DM with ophthalmic complications, 0.978 for T1DM with renal complications, 0.197 for T1DM with peripheral circulatory complications, and 0.345 for T1DM with neurological complications. No evidence of horizontal pleiotropy or heterogeneity was found in these MR analyses. (Figure 2; Supplementary Table S5).

### 2. Causal effects of T2DM on OM

Using IVs for T2DM, we found evidence linking genetically predicted T2DM to increased OM risk (primary dataset: IVW: OR = 1.13, 95% CI 1.056–1.209, *p =* 4.20E-04; validation dataset: IVW: OR = 1.317, 95% CI 1.14–1.522, *p =* 0.007; Meta-analysis: OR 1.203; 95% CI 1.038–1.395; p=1.85E-04) (Figure 2). Furthermore, after Bonferroni correction, the results remained statistically significant. The three MR methods showed consistent directions. After MVMR analysis controlling for BMI, statistical significance remained between T2DM and OM (Figure 3; Supplementary Table S7). No outliers were detected by MR-PRESSO, and no heterogeneity was observed by Cochran’s Q test. MR-Egger intercept tests found no horizontal pleiotropy. Leave-one-out analysis showed consistent T2DM results (Fig. 2). Leave-one-out and scatter plots are provided in Supplementary Figures 5-8.

**Figure. 3.**
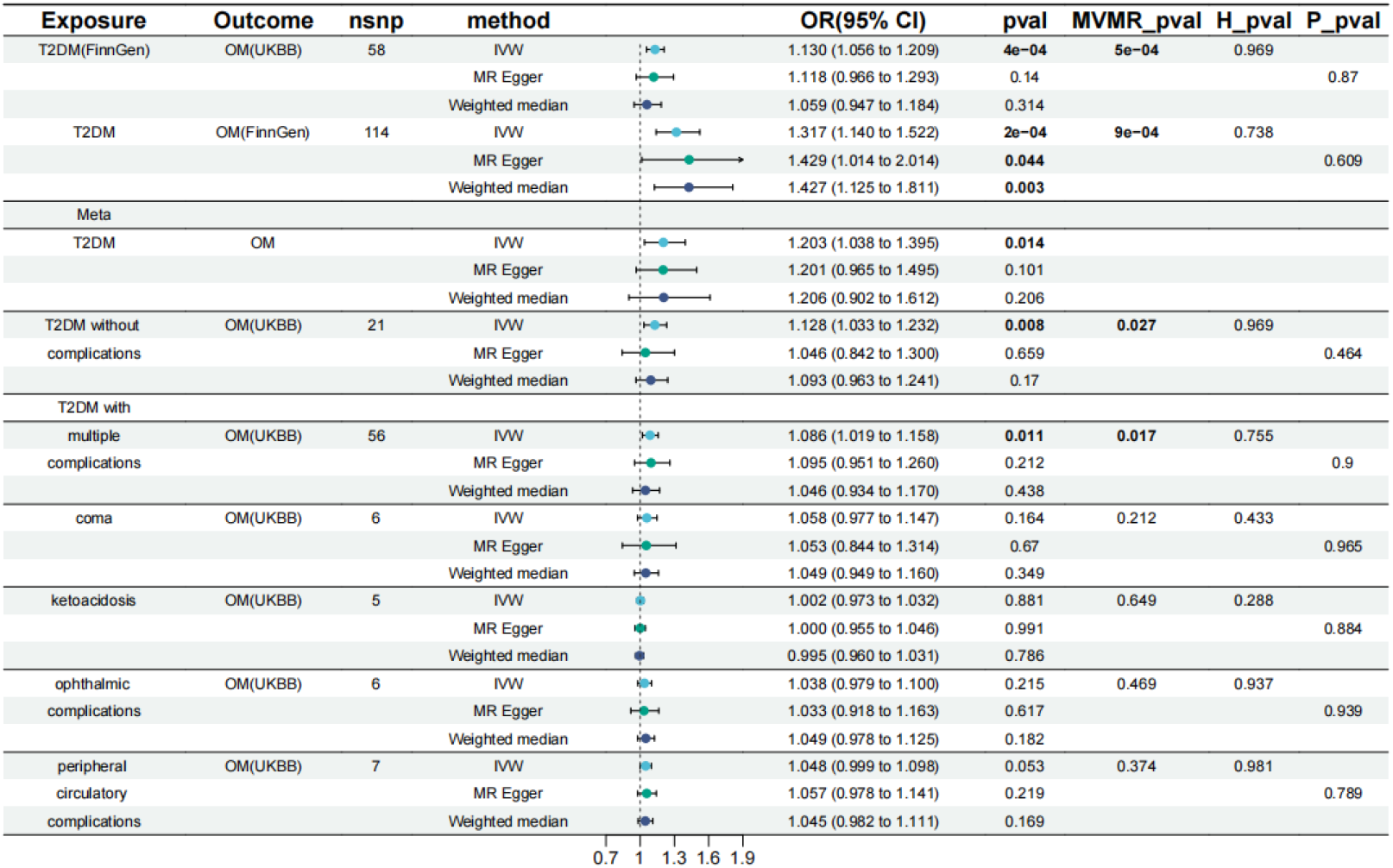
Genetically predicted type 2 diabetes and its complications: associations with osteomyelitis. T2DM, type 2 diabetes mellitus; OM, osteomyelitis; UVMR, Univariable MR; MVMR, multivariable mendelian randomization; OR, odds ratio; CI, confidence interval; UKBB, United Kingdom Biobank; IVW, Inverse Variance Weighted; H, Heterogeneity; P, Pleiotropy. The IVW p-value (<L0.05) was considered a statistically suggested difference. IVW p-values below the Bonferroni-corrected threshold were considered statistically significant.

Using GWAS data for T2DM complications from the FinnGen database, we initially observed suggested causal correlations between both T2DM without complications and T2DM with multiple complications and OM (IVW: OR 1.128; 95% CI 1.033–1.232; p□=□0.008; OR 1.086; 95% CI 1.109–1.158; p□=□0.011), respectively. However, after multiple testing correction, these causal correlations disappeared. Furthermore, analyses for T2DM with other complications revealed no significant causal correlations with OM. Specifically, IVW-P values for OM were 0.164 for T2DM with coma, 0.349 for T2DM with ketoacidosis, 0.215 for T2DM with ophthalmic complications, and 0.053 for T2DM with peripheral circulatory complications. These MR analyses showed no evidence of horizontal pleiotropy or heterogeneity (Figure 3; Supplementary Table S9). Insufficient genetic instruments were available for MR analyses of other T2DM-related complications.

The findings suggest that genetically predicted T2DM, rather than T2DM with complications, is a risk factor for OM.

### 3. Inconsistent Causal Effects of Glycemic Traits on OM

There was no causal relationship between fasting insulin and OM in either the primary dataset (IVW: *p =* 0.566) or the validation dataset (IVW: *p =* 0.463). However, fasting glucose showed inconsistent effects on OM. In the primary dataset, the IVW analysis yielded a p-value greater than the threshold (IVW: OR = 1.265, 95% CI: 0.972-1.645, *p =* 0.08), while in the validation dataset, the p-value was smaller than the Bonferroni-corrected threshold (IVW: OR = 2.347, 95% CI: 1.276-4.318, *p =* 0.006). However, meta-analysis did not support fasting glucose as a risk factor for OM (IVW: *p =* 0.11). Similarly, inconsistent results were observed for HbA1c levels and OM in the primary dataset (*p =* 0.29) and the validation dataset (IVW: *p =* 0.04), with the latter failing the Bonferroni correction for multiple testing. Nevertheless, meta-analysis indicated that HbA1c levels were not associated with an increased risk of OM (IVW: *p =* 0.59). Sensitivity analyses confirmed no evidence of heterogeneity or horizontal pleiotropy (Table 1, Supplementary Tables S11, S13 and S15). These findings suggest that there may be complex relationships between glycemic traits and OM.

**Table 1.**
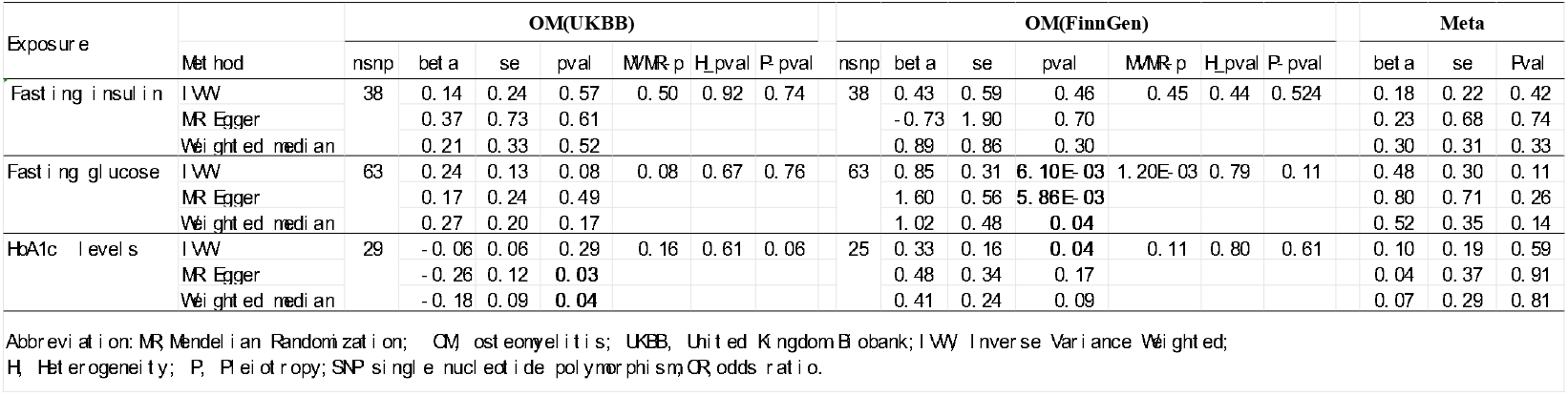
MR and Sensitivity alysis of Glycemic Traits with the risk of OM.

### 4. T2DM mediates the causal effect of RA on OM

We employed two-step mediation MR analyses in both primary and validation datasets to explore T2DM’s mediation between RA, and OM. RA was associated with increased OM risk (IVW: OR□=□1.058, 95% CI 1.005–1.114, *P*LJ*=*□0.031), with a 25.8% mediation effect (95% CI 9.0%–42.5%) in the primary dataset. Consistent results were observed in the validation dataset analysis, with T2DM mediating 14.5% (95% CI 6.1%–23.0%) of the RA-OM relationship. No heterogeneity or horizontal pleiotropy was detected in any analysis (Table 2; Supplementary Table 17).

**Table 2.**
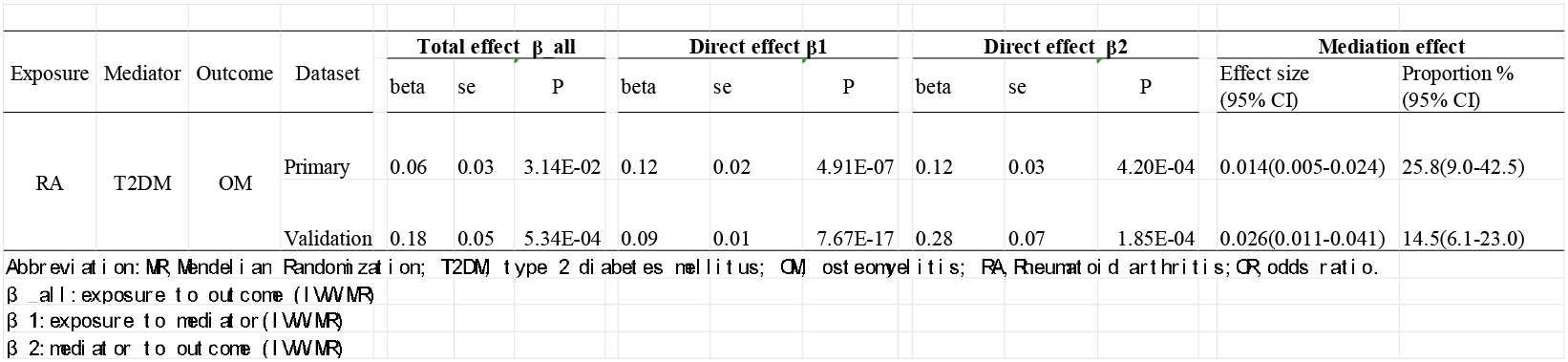
Mediation effect of T2DM in the association between RA and OM.

| Exposure | Mediator | Outcome | Dataset | Total effect $\beta_{all}$ | | | Direct effect $\beta_1$ | | | Direct effect $\beta_2$ | | | Mediation effect | |
| --- | --- | --- | --- | --- | --- | --- | --- | --- | --- | --- | --- | --- | --- | --- |
|  |  |  |  | beta | se | P | beta | se | P | beta | se | P | Effect size (95% CI) | Proportion % (95% CI) |
| RA | T2DM | OM | Primary | 0.06 | 0.03 | 3.14E-02 | 0.12 | 0.02 | 4.91E-07 | 0.12 | 0.03 | 4.20E-04 | 0.014(0.005-0.024) | 25.8(9.0-42.5) |
|  |  |  | Validation | 0.18 | 0.05 | 5.34E-04 | 0.09 | 0.01 | 7.67E-17 | 0.28 | 0.07 | 1.85E-04 | 0.026(0.011-0.041) | 14.5(6.1-23.0) |

Abbreviation: MR, Mendelian Randomization; T2DM, type 2 diabetes mellitus; OM, osteomyelitis; RA, Rheumatoid arthritis; OR, odds ratio. $\beta_{all}$ : exposure to outcome (IVW) $\beta_1$ : exposure to mediator (IVW) $\beta_2$ : mediator to outcome (IVW)

## Discussion

Our MR study revealed distinct causal effects of T1DM and T2DM on OM risk. Specifically, T2DM showed a significant association with increased OM risk, while T1DM and its complications did not exhibit a causal relationship with OM. Additionally, glycemic traits (fasting insulin, fasting glucose, and HbA1c) showed inconsistent effects on OM. Furthermore, we identified that T2DM mediates the causal effect of RA on OM. These findings deepen our understanding of the differing roles of T1DM and T2DM in OM development and provide avenues for further mechanistic investigation.

The discovery that that T1DM and its complications did not exhibit a causal association with OM challenges our conventional understanding and contradicts some previous study findings. ^27–29^. Traditionally, diabetes, including both T1DM and T2DM, has been linked to an elevated risk of various infection complications, such as OM. Moreover, individuals with diabetes face a higher mortality risk from infections compared to the general population, with T1DM patients particularly susceptible^30^. However, our Mendelian randomization analysis, which provides more robust causal inference compared to observational studies, suggests otherwise. This unexpected result prompts a reevaluation of the assumed causal relationship between T1DM and OM. One possible interpretation could involve the complex interplay between diabetes subtypes and their respective pathophysiological mechanisms. While T2DM is commonly associated with metabolic abnormalities and chronic inflammation, the etiology of T1DM involves autoimmune destruction of pancreatic beta cells. It is plausible that the mechanisms underlying T1DM may not directly contribute to the development of OM, as observed in our study. Another explanation for this discrepancy could be the differences in study design and inherent biases between observational studies and Mendelian randomization. Observational studies are limited to establishing associations and are susceptible to confounding factors, reverse causation, and measurement errors, which may lead to spurious associations. In contrast, Mendelian randomization leverages genetic variants as instrumental variables to mimic the random assignment of exposures, thereby reducing bias and providing more reliable causal estimates^31^.

The finding that T2DM is significantly associated with increased OM risk aligns with previous clinical studies^1,5,6^. However, our analysis reveals an unexpected causal relationship between glycemic traits and OM risk. The absence of associations with fasting insulin and HbA1c implies that insulin and HbA1c levels may not contribute to OM risk. Inconsistent causal effects of fasting glucose were observed between the primary dataset and validation dataset, and meta-analysis did not support fasting glucose as a risk factor for OM. Therefore, T2DM may influence OM risk through alternative mechanisms beyond high blood sugar, such as immune response dysfunction ^32^ or metabolic disturbances ^33^. These findings emphasize the need for further research to elucidate the specific pathways through which T2DM affects OM risk. Additionally, they underscore the importance of comprehensive risk assessment in diabetic patients, beyond glycemic control alone.

Previous research has established RA as a common risk factor for both T2DM and OM^7,9,10^. Our study further investigated their associations with T2DM and OM risk, revealing the extent to which T2DM mediates RA’s causal effect on OM. These findings underscore the importance of addressing RA to prevent OM and highlight T2DM’s role in connecting RA with OM. Such insights have significant clinical implications, suggesting targeted interventions for diabetic patients with RA to mitigate their heightened susceptibility to OM, particularly those requiring orthopedic procedures like fracture fixation or joint replacement. Strategies focusing on optimizing glycemic control and managing inflammation in this patient population may help reduce the risk of OM.

Our study reveals distinct causal effects of T1DM and T2DM on OM risk, underscoring the need for tailored disease management. Further validation is essential for a comprehensive understanding of these findings. The observed differences in OM risk between T1DM and T2DM warrant further investigation into underlying pathophysiological heterogeneity. Future research should focus on identifying biomarkers to stratify diabetic patients based on their susceptibility to OM and related complications. Additionally, our findings suggest potential drug repurposing for OM management among diabetic patients, emphasizing the importance of clinical trials to validate efficacy and safety profiles. While our study provides valuable insights, prospective clinical studies are needed to validate our findings and improve clinical outcomes for diabetic patients.

Our study boasts several strengths. Firstly, it pioneers the investigation into the causal link between DM and OM using MR approaches. Notably, it marks the inaugural documentation of the absence of a causal effect of T1DM on OM. Secondly, we employ MR techniques to mitigate potential confounding biases and ensure reliable causal inferences. Thirdly, our use of GWAS data primarily from European ancestry populations minimizes population stratification bias. all outcomes underwent rigorous validation through various sensitivity analyses, including inverse-variance weighted (IVW), MR-Egger, MR-PRESSO, leave-one-out analysis, and MVMR, with multiple testing corrections applied. Furthermore, consistent results across different datasets and meta-analysis underscored the robustness of our findings.

Our study has limitations affecting generalizability and interpretation. Focusing on a single ethnic group restricts broader applicability^34^. Replicating in diverse cohorts is necessary. Reliance on summary statistics from GWAS may introduce misclassification bias. Additionally, while benefiting from large-scale data, our sample size may be insufficient to detect small effect sizes or rare variants. Potential pleiotropy in Mendelian randomization analysis and residual confounding remains concerns, despite sensitivity analyses.

**In conclusion**, our study offers compelling evidence of a significant causal relationship between genetically determined T2DM and heightened risk of OM, while T1DM shows no causal effects. Additionally, our findings highlight the role of T2DM in mediating the association between RA and OM. These insights hold promise for improving clinical outcomes among diabetic individuals and those at risk of OM.

## Data Availability

All data produced in the present study are available upon reasonable request to the authors

https://www.finngen.fi/en/access_results

https://gwas.mrcieu.ac.uk/

https://www.ebi.ac.uk/gwas/home

## Acknowledgments

We want to acknowledge the participants and investigators of the FinnGen study, UK Biobank, and other GWASs included in this study. Without the dedication of these organizations and their members, this article would have been difficult to complete.

## Data Availability Statement

All data generated or analyzed during this study are included in this article and its supplementary information files.

## Ethics Statement

Not applicable

## Author Contributions

L.T. conceived the project, designed the study and revised the manuscript.; YH.L. wrote the manuscript; CY.Z. collected data from the public database; L.T.,YH.L. and LM.Y. performed the study, and all authors reviewed the manuscript.

## Consent for Publication

Not applicable

## Abbreviations

T1DM: type 1 diabetes mellitus
T2DM: type 2 diabetes mellitus
OM: osteomyelitis
IVW: Inverse variance weighted
IVs: Instrumental variables
SNPs: Single Nucleotide Polymorphisms
UVMR: Univariable MR
MVMR: Multivariable MR
OR: Odds ratio
CI: confidence interval
GWAS: Genome-Wide Association Study
UKBB: United Kingdom Biobank

